# Barriers and Perceived Facilitators for PrEP Use Among Women of Color in South Florida: Findings from the *Empowering Women to Take Control of Their Sexual Health Summit*

**DOI:** 10.1101/2023.05.23.23290385

**Authors:** Elena Cyrus, Karina Villalba, Michele Jean-Gilles, Rhonda Rosenberg, Amanda Ichite, Evelyn Ullah, Evelyn Lovera, Aryah Lester, Sandra Neptune, Hannah Wilson, Maude Exantus, Brenda Lerner, Robert Cook, Jessy G. Dévieux

## Abstract

This paper aims to summarize conference proceedings and testimonies from the “Empowering Women to Take Control of Their Sexual Health Summit” convened in South Florida. In the present study, the phrase “women of color” refers to 279 cisgender and transgender African American, Latina, and Haitian women. Data were collected through three 90-minute group discussions conducted at the conference Individual-, social-, and structural-level PrEP barriers and facilitators were identified. Individual level barriers included: medical distrust, limited knowledge about PrEP, and economic dependence on sexual partners. Participants discussed cultural gender norms and roles as overarching social barriers, with religiosity reinforcing these norms. Structural barriers included: health literacy, health insurance coverage, immigration status, and structural racism. Community attributes that acted as facilitators of PrEP use were resilience and adaptability. Increasing PrEP utilization among women of color requires a multi-tiered approach to comprehensively address structural and community level barriers.

## I. BACKGROUND

HIV continues to be a significant health risk for women in the United States (US), particularly among minoritized women in the South. For example, women of color in South Florida represent 88% of new HIV cases among all women, but compose only 67% of all women in the region (Florida Department of Health, 2022). Although new HIV cases among minority women in South Florida have remained unchanged since 2014 (18% of all cases), these data sharply contrast the downward trends among all other heterosexual subgroups reported in recent years (Florida Department of Health, 2022). Furthermore, for minoritized women living in South Florida, risks extend beyond individual risk factors to environmental risks, such as living in a high prevalence area (Patel et al., 2019), because between 2009 and 2014, the largest proportion of new HIV cases and people living with HIV/AIDS in the US resided in the Southeast (Centers for Disease Control and Prevention, 2018).

PrEP is an effective self-administered biomedical tool that can empower women to prevent HIV transmission autonomously; yet, in the US, it is underutilized among women and a major challenge is the lack of awareness about PrEP, particularly among minoritized women and their healthcare providers (Braksmajer et al., 2016). To improve awareness of PrEP that could spark action among individuals and community agencies in South Florida, the summit was convened in 2018 with a focus on empowering women to take control of their sexual health through knowledge of biomedical HIV prevention methods such as PrEP.

PrEP was approved by the Food and Drug Administration (FDA) in 2012, with initial indication for high-risk groups such as men who have sex with men (MSM) (Grant et al., 2010). In 2018, the CDC expanded this indication to include women and adolescents (Centers for Disease Control and Prevention, 2018). PrEP use has increased among MSM and may be contributing to declines in HIV rates in certain US regions (Chan et al., 2018; de Mendoza, 2017; Stahlman et al., 2017); however, utilization among women remains low, particularly among minoritized women who can be at greater risk for HIV infection (Ravindran et al., 2020). At an HIV clinic in New York City, compared to cisgender male patients, cisgender women had the lowest PrEP use (2.1% vs. 60.1%) (U. Belkind, 2018). In 2017, in the State of Florida, AIDSvue estimated 4/100,000 women used PrEP versus 83/1000 men (Sullivan PS, 2020). A study among transgender women in Florida supported statewide findings that PrEP awareness was high (65%); however, there was very low active use (8.2%) (Holder et al., 2019).

The primary objectives of the summit were to (a) gather and connect key community stakeholders and providers to discuss the landscape of PrEP in local communities and provide updates on available resources; (b) organize group discussions to identify PrEP use barriers and facilitators among racial/ethnic minoritized women in South Florida; and (c) generate recommendations to effectively tackle intersection factors hindering PrEP use among women.

The present article reports on the Summit’s exploratory “breakout” sessions, which functioned as focus groups and explored African American, Latina, and Haitian women’s use and knowledge of PrEP in South Florida. To our knowledge, no previous discussion on factors preventing and/or facilitating PrEP use among women of color in South Florida has been presented in the literature, and the findings in this paper provide a wealth of information engendered at this progressive summit.

## II. METHODS

On May 4, 2018, Florida International University (Miami, FL) hosted the *Empowering Women to Take Control of their Sexual Health Summit*, to address the use of PrEP among women. Attendees (n=279) included underrepresented racial/ethnic minority cisgender women and transgender women who were patients, activists, healthcare providers, researchers, and lay community members representing different sectors of the HIV community in Florida. A plenary session provided information on the availability of biomedical tools to end HIV transmission [i.e.., PrEP and post exposure prophylaxis (PEP)] and breakout sessions (hereafter referred to as *focus groups*), were conducted to understand perspectives and realities of African American, Latina, and Haitian women in South Florida.

Each focus group (n=3) explored the experiences of a racial/ethnic minority community: African American, Latina, and Haitian. Each group was led by two moderators who served as discussion facilitators and note takers. The two facilitators included an academic researcher and a lay member from the community. Each group included 70-80 participants and lasted approximately 75 minutes.

A group discussion guide was developed to systematically explore issues related to the participants’ perceived barriers and facilitators to PrEP. Discussions converged on topics related to stigma, social justice movements and intersectionality, the power of messaging and social media, and the importance of storytelling to support positive social norms, culture, and tradition in the women’s empowerment movement. To understand how to improve access and engagement in care, discussions explored participants’ perceptions of women’s strengths (e.g., resilience, adaptability and flexibility, and their caregiver role within their families) in the context of South Florida.

The participant responses were documented without any personally identifiable information. Notes were summarized at the time of the group session. Before the end of each focus group discussion, annotated summaries were reported back to the group for verification and subsequently presented to the wider conference audience at the Summit’s closing plenary session. The notes and summaries were compiled, transcribed, and analyzed by the authors. Salient and recurrent themes from each session are reported. Barriers and facilitators, whether perceived or actual, can occur at multiple levels (i.e., individual, interpersonal, community, and structural levels) (Philbin et al., 2016; Wingood et al., 2013); therefore, we present findings from the focus groups conducted at the Summit within the social ecological framework (Figure 1) to comprehensively explore factors influencing PrEP use. Focus group discussions and related processes were submitted to the FIU Institutional Review Board (IRB) and were found to be exempt from human subjects’ research protocols.

**Figure 1.**
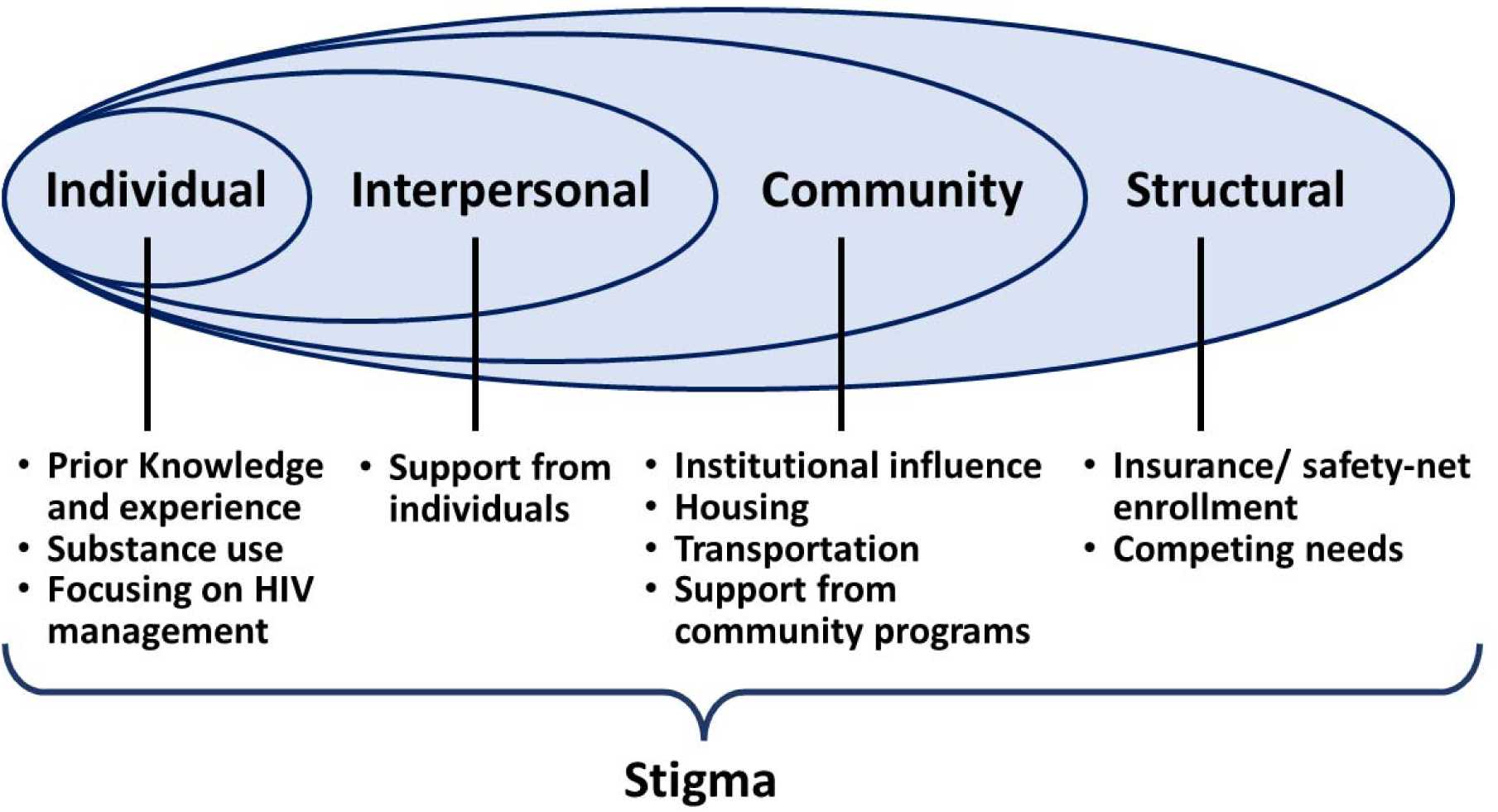
Description: Social ecological framework illustrating women-specific PrEP barriers and facilitators.

## III. RESULTS

The discussions on African American, Latina, and Haitian women’s use of PrEP ranged widely, reflecting the diverse backgrounds, expertise, and experiences of the participants and the specific reality of each group. There was congruence among the participants on systemic factors preventing PrEP use; however, at the community or individual level, more contextual and culturally specific barriers and recommendations were discussed.

### Barriers to PrEP

#### a. Structural and systemic barriers

Participants in all the groups highlighted significant problems with discrimination and inequity in healthcare systems. There was discussion on US federal politics and policy impacting overall access to care in local health systems and differential impact among communities or populations. For example, immigration status represents a layer of perceived discrimination within the healthcare system, with negative implications for unauthorized individuals. In the Haitian focus group, this immigration-related barrier was cited as a reason why women are not exposed to mainstream public health messaging and may not be aware of community resources available to them for HIV testing and prevention. Several participants acknowledged the lack of trust in government entities and publicly funded health care services. One participant stated, “undocumented immigrants don’t trust doctors or clinics; they are afraid they will end up being deported.”

Healthcare environments (e.g., clinics, private medical offices, hospitals, etc.) were noted as being implicitly biased because they were perceived as being part of or representing systems that were established and influenced by sexist or racist dogma that can manifest as barriers to HIV prevention and other services. In the African American group, participants noted the lack of representation of people of color in clinical trials and campaigns, which helped form their general impression that existing programs and policies were not reliable, relevant, or safe for their communities. Participants also shared that African American and Latina women experienced challenges related to health insurance coverage, time constraints with regard to attending multiple doctors’ visit that might be required if they are on PrEP, and health care costs when they considered accessing PrEP. Participants were concerned about the lack of health insurance and unaffordable PrEP prices, as well as doctors who are not willing to prescribe PrEP due to their own biases and stigma around people who use PrEP who can be considered vulnerable or at increased risk for HIV transmission. There was consensus that the existing profit motivated health care model in the US is a barrier (Galvani et al., 2020), where the individual’s overall wellbeing is not necessarily prioritized by healthcare providers because the provider’s profit margin is prioritized over patient health.

African American and Haitian women discussed a disconnect with—and lack of respect from—healthcare providers. Latina women identified similar issues relating to providers’ attitudes and biases acting as PrEP barriers. Among the Black ethnic groups (i.e., African American and Haitian women), women felt that structural racism and discrimination led to chronic social stress and stemmed one’s desire to access healthcare. On the later stage of the care continuum, there were reported disparities around prescription of PrEP (Calabrese et al., 2014).

Five Latina and African American transgender women contributed to the focus group discussions. Barriers for transgender women included the struggle to balance their primary health care and prevention needs with competing personal priorities such as gender affirmation procedures (e.g., hormone therapy vs. PrEP), or other basic needs such as housing, food security, and transportation. Transgender women may have difficulty prioritizing HIV prevention or overall health in the face of extreme discrimination and marginalization, which limits their opportunities for upward social mobility and consequently their healthcare engagement.

#### b. Intrapersonal and Community Factors: social barriers

At the intrapersonal level, four common themes emerged among the groups: (1) cultural norms and beliefs, (2) gender roles, (3) religion, (4) stigma around sexual health and sexuality. *Machismo*, reported by participants in the Latina group, paralleled the concepts of misogyny or chauvinism discussed among African American and Haitian women. The women discussed cultural norms rooted in tradition, as well as religiosity promoting machismo/misogyny and/or chauvinism, which deterred women from considering, discussing, or electing to use PrEP. PreP use may not be supported by their sexual partners within the context of a relationship or adherence to social norms or religious beliefs.

African American women explained the differences between public and private personae and norms positing that Black women are stereotyped as aggressive or ‘strong’ in public spaces but lacked power in the home. This concept was attributed to the influence of culturally ascribed attitudes and behaviors shaping gender roles in their communities. Participants in the Haitian group noted that Haitian women faced similar cultural challenges with men often having more decision-making power, in terms of sexual health issues, within a relationship. Sexual and reproductive health decisions included condom use, screening for HIV and/or sexually transmitted infections (STIs), seeking treatment options, etc. Women stated it is not uncommon for a sexual partner to undermine or ignore women’s desires regarding their sexual and reproductive health.

Culturally ascribed gender roles were also identified in the Latina focus group as a factor preventing women’s use of HIV risk reduction options. Among Latinas, family traditions and gender roles also shape attitudes and beliefs about sexual education which may not be centered on women’s health needs, which can in some cases increase HIV/STI risk. One participant stated, *for women to be open to use PrEP, they needed to change their beliefs and attitudes about sex and their roles in the family*.

Shifting cultural roles toward the evolution of gender norms is a challenge for minoritized women as well as their health providers who are not necessarily trained in culturally relevant or gender salient subjects that impact their healthcare. Furthermore, Latino families often adhere to gender roles and are invested in maintaining traditional values across generations (Miville, 2017). For example, cultural values such as *familismo* and *marianismo* place Latina women in a submissive role—pressuring them to prioritize family needs before individual personal interests (Castillo et al., 2010; Finch & Vega, 2003). Cultural expectations influence gender role ideologies, sexual identity, and power imbalance by promoting women’s sexual silence (Galanti, 2003; Ibañez et al., 2017; Villarruel, 1998). Among Latina women, sexual silence perpetuates sexual coercion and inhibits women from discussing sexual and reproductive health issues (e.g., condom use, extramarital relationships) with their partners (Villar-Loubet et al., 2011). Women in the Latina focus group reaffirmed that a major barrier to PrEP use is the lack of conversation about sex in the home.

According to the Latina participants, religion plays a determining role in sexual health, sexual practices, and use of condoms and other HIV prevention tools. These views are supported by findings in previous research (De Jesus, 2016). Research suggests that religiosity can support traditional Latina attitudes on sexuality, thereby inhibiting sexual freedom or conversations about sexuality (De Jesus, 2016).

Before adulthood, Latino parents shy away from discussing sexual topics regarding their children’s sexual orientation and/or their sexuality (Guzman et al., 2013). The lack of information regarding sexual and reproductive health in Latina homes is compounded by the dearth of accurate or current sexual education in Florida school systems. Florida is one of 23 states that require schools to include sex education and HIV prevention classes in the curriculum; however, the focus group participants in this study expressed reservations about the sexual education curriculum being completely evidence based or comprehensive.

Prior studies have demonstrated that sex education programs in Florida’s public schools vary widely in content and duration and implementation with some being more robust than others (Caroline Wheeler-Hollis, 2019; Megan Reeves, 2019; National Conference of State Legislatures, 2020).

Participants in the Haitian focus group also noted the influence of religion and traditional gender roles on sexuality. Haitian churches can have influence within the community, perpetuating strong misogynistic or chauvinistic messages “that women should obey their husbands” (Caroline Wheeler-Hollis, 2019; Crystelle Lee, 2020). Several women talked about how some churches condone acts of infidelity by husbands, thereby placing their wives at increased risk for HIV/STI acquisition.

Haitian focus group participants discussed the influence of spiritual leaders on sexual health decision making among families in the congregations. Examples were given of some church leaders advising people living with HIV against treatment because *Jesus will save them.* The religious community was perceived as a significant barrier to truthful sex education and preventing HIV and group members indicated that, in some cases, religious leaders were perpetrators or enablers of sexual abuse and sexual harassment. The suppression of women’s roles through religion or other cultural norms prevents open discussions of sex and associated risk, and also disempowers women.

African American participants discussed the role of stigma and discrimination that influenced the tone and direction of their medical encounters or visits with their doctors. Participants—particularly those living with HIV—expressed concerns that personal sexual behaviors were potential sources of stigma and judgement from providers and family members. Women stated that this perceived judgment made them less confident during a medical encounter to be candid with their providers, making it more challenging for women to screen or disclose their HIV status and/or access treatment and care.

Previous studies have demonstrated that stigma hinders minoritized women’s ability to disclose their status and seek treatment; consequently, many women forgo testing, preferring ignorance of their status rather than potentially exposing themselves to exacerbated HIV-related stigma and discrimination (Frank et al., 1985).

#### c. Individual barriers

During the Summit, women cited their distrust of providers, health literacy level, and economic status as determining factors that informed their awareness of PrEP as a prevention option; these factors also informed their perception of how easy it is to access PrEP.

Black women expressed distrust in the scientific community and had concerns about trusting providers. Some women felt that providers may not protect their privacy or prescribe PrEP without judgement, which motivated them to seek medical care externally due to fears of stigma or a confidentiality breach. Poor health literacy or understanding of the health system exacerbated this distrust, which was compounded by their perception that providers in their local communities did not always ask relevant questions. Low health literacy contributed to PrEP misinformation being spread within their communities. Concerns were voiced about women being vulnerable to “charlatans” or HIV deniers that spread false information about HIV that is not evidence-based, including unsubstantiated cures for HIV (Garett & Young, 2022; Grov et al., 2021). For example, cases of non-approved treatment or unlicensed providers, such as the use of *Picuristes* (i.e., lay injectionists) among Haitians in Miami-Dade County, potentially increased women’s vulnerability and risk (Rahill & Mallow, 2011).

Women’s economic level was also identified as an individual barrier across all the groups. There was discourse about women’s economic dependency on their partners, which disempowered them and negatively impacted their vulnerability and sexual risk. Having a low income is a well-documented barrier for accessing care but, for many women, economic dependency may also prevent them from adopting HIV reduction strategies or making independent decisions about their sexual health.

Participants stated there was a lack of opportunity to have open conversations with their partners about sex and sexual health and in the home, which was reinforced by prevailing cultural norms.

### Facilitators of PrEP use

There was a strong recommendation from all groups to mobilize communities through active and strong engagement of community leaders and community-based organizations that align with the needs and desires of their communities. One woman in the African American focus group stated: *there are those without fear and passion to work in communities—[we] need to reactivate*! African American and Haitian women drew from their communities’ history of activism as strengths that should empower and facilitate access to healthcare. The legacy of the civil rights movement, among other formidable contemporary social movements (Fernando Calderón, 1992; PETER DREIER, 2020), has survived and shaped their communities; this legacy can be applied to the issue of women’s empowerment and prioritizing sexual health (Byrd, 2015; CHARLES M. PAYNE, 2003; Wah et al., 2004). The participants described the importance of tapping into the sense of historical pride, civic duty, and responsibility because these align well with messages of HIV prevention, including PrEP use. Citing additional community strengths, African American women discussed the ability of communities to convene and create power in numbers, and they discussed the resilience and strength that is developed through family structures (Gwadz et al., 2021; Koch et al., 2022).

Haitian women noted their respect for health care providers, respect for education, and interest in health and healthy living as community strengths and as resources that could be used to expand campaigns and prevention strategies. Although HIV prevention strategies often leverage these community assets, these efforts can be undermined by the antithetical cultural and social barriers previously noted.

Latina participants offered the possibility of creating awareness about PrEP through personal testimonies from community members already using PrEP. One participant recounted a personal experience of being open about their HIV status to advocate for people living with HIV: *I have HIV and to bring awareness about my disease, I decided to wear a t-shirt that said: ‘this is what HIV looks like*. The focus group agreed this was a powerful strategy that could also be used for PrEP by way of role-modeling positive and healthy behaviors associated with PrEP use.

### Strategies and recommendations to promoting linkage to PrEP care and PrEP use

Participant recommendations for improving access and use of PrEP (1) highlighted the need for implementing patient-centered care models, (2) addressing the cultural diversity of all women, and (3) involving the community as an equitable partner to support the empowerment of women especially around their health care decisions.

Specific recommendations were:

#### 1. Educate healthcare providers to promote and support prescription of PrEP

Tacking misinformation or lack of information by providing equitable education and dissemination of PrEP information among healthcare providers in all segments of society were common themes. A strategy was recommended to create a more robust multicultural, diverse medical workforce equipped and trained to serve all communities, who are willing to develop or employ existing strategies that effectively link individuals to PrEP services.

Another consistent theme was that awareness and knowledge about PrEP among providers needed to be improved to address disparities, and to ensure that PrEP is prescribed without judgement, equitably, among those who are eligible and decide to use PrEP. Determining current levels of knowledge among medical providers is a possible point of intervention to expand PrEP prescriptions and referrals, as provider knowledge of PrEP has been correlated with future willingness to prescribe (Adams & Balderson, 2016). Studies have suggested that many specialized providers are aware of PrEP and support its use as a public health intervention, though knowledge and acceptance of PrEP are usually lower among general doctors (Krakower & Mayer, 2016).

Qualitative studies have shown that some providers did not perceive themselves to be well-positioned to prescribe PrEP because they do not serve populations considered to be at increased risk to acquire HIV (Krakower et al., 2014; Laws et al., 2014). Some primary care doctors believed that they did not have relevant experience to prescribe medications such as PrEP (Krakower et al., 2014). Additionally, concerns about the efficacy and long-term safety of PrEP could limit prescribing behaviors and create resistance around performing routine HIV risk assessments needed to recommend PrEP safely to patients (Krakower & Mayer, 2016). Thus, strategies needed for PrEP provision should engage a broader array of healthcare providers beyond primary care physicians. As suggested in the Latina focus group, the scope for educating health professionals should be expanded into medical schools, with a suggestion from that group to *develop a policy that requires medical schools to teach medical providers about cultural awareness and sexual health*.

Ultimately to increase PrEP prescriptions, participants called for the development and implementation of effective educational interventions to help providers acquire the knowledge and skills to screen patients and consistently record sexual history at intake to determine HIV risk and those who are eligible for PREP (Adams & Balderson, 2016). In response to this recommendation, the Summit organizers secured NIH funding that supported the development of education videos that are currently being pilot-tested and prepared for dissemination with collaborating community agencies.

#### 2. Address fears among immigrant communities

Public health campaigns that address fears related to immigration status were a key recommendation. Several participants acknowledged the lack of trust immigrant communities feel toward government entities and public health providers. Forgoing treatment for chronic conditions and preventive medical care has been widely reported by immigrant groups following the 2018 changes to the “public charge” rule, which is a policy that determines if a person can be denied a green card, visa, or admission into the United States and also requires those seeking permanent and legal status to prove they will not be a burden to the US (Bernstein et al., 2019).

The Florida Department of Health in Miami-Dade County has maintained outreach programs to provide PrEP information and medical assistance with the provision of subsidized PrEP statewide for uninsured individuals (Human Rights Campaign, 2018). While these efforts by the state are crucial, more targeted messages are needed to address the unique fears of deportation and respond to the Federal policies that have challenged access to care among immigrants, both unauthorized individuals and non-citizens.

#### 3. Promote patient-centered appropriate health services

Developing trust between patients and providers, by employing patient-centered culturally tailored care models, can facilitate provider-patient communication and promote patient empowerment in minority racial/ethnic groups. These models embrace the cultural diversity of the patients, in contrast to stereotypical views that can undermine patient-provider trust (Tucker et al., 2011). For Haitian communities, there was a suggestion that during a medical encounter, providers should assist patients with their personal priorities and immediate medical concerns to develop rapport before attempting to solicit sensitive information from them that may be required for the medical history and intake. For example, before talking about sexual behavior to assess risk, participants may be more concerned with fertility issues, or gender affirmation procedures if they are transgender, and will prioritize these issues. Furthermore, providers should exercise caution when initiating patient education, considering those who need more time to process new information and concepts.

Culturally tailored approaches should be inclusive of traditional healing systems (Andrews et al., 2013), and Haitian women demanded culturally diverse medical providers to empower community members to be better self-advocates to obtain optimal healthcare. Compared to non-Latino White patients, medical visits between HIV providers and Latino patients tend to be less patient-centered with less discussion about psychosocial issues (Morales-Aleman & Sutton, 2014). Less communication with patients of color may be because providers are disconnected from patients’ cultural context and are unprepared to explore unfamiliar psychosocial domains (Valdez et al., 2011). Alternatively, because of cultural norms, Latina women do not consider themselves to be well-positioned to be transparent when discussing sexual topics with their doctors (Morales-Aleman & Sutton, 2014). Several Latinas at the Summit acknowledge the cultural norm of seeking approval and validation from their doctors, which might prevent them from disclosing sexual history to their medical providers or request information about how to prevent HIV/STIs.

#### 4. Culture-centered education and dissemination of PrEP information

Haitian and Latina focus groups discussed engaging people with political power and key stakeholders or ‘community influencers’ to disseminate accurate community-wide HIV prevention and PrEP messages. These community influencers can be used to (a) endorse the promotion of sexual health education, including HIV/AIDS, and decrease stigma around HIV/AIDS and prevention methods in public schools; (b) obtain support from churches and educate spiritual leaders on the benefits and necessity of prevention and treatment; and (c) advocate for women’s rights to protect themselves and receive treatment.

Summit attendees also highlighted the importance of reaching the community through proven community-specific communication mediums to increase awareness of HIV prevention and PrEP. For example, social media strategies were highlighted as viable methods to reach unauthorized immigrants. Another recommendation was to make health information available to the Haitian and Latino communities through local radio stations. Radio is useful for dissemination among those who may be illiterate or who do not speak English. Similarly, the Latina focus group suggested that increasing PrEP awareness through popular Spanish soap operas or *Telenovelas*. Telenovelas can be used by health communication experts to create a thematic storyline that weaves PrEP and HIV prevention into the main plots. The telenovela, which already has an expansive audience in the Latino community (Castaneda et al., 2013), can be a vehicle for PrEP awareness and dissemination to Latino audiences in the US and globally.

An additional strategy considered by the Latina focus group included the promotion of candid discussions with their children about sexuality including HIV/STI risk. Programs that were discussed and recommended for the Latino/Hispanic community included using evidence-based programs such as *Back to Basics* (Germain et al., 2009), *Saber es Poder* (Greather than AIDS, 2022), Getting to Zero (Florida Department of Health, 2017), and Undetectable Equals Untransmittable (U=U) Campaign (Calabrese et al., 2021; Cohen, 2018).

Another important recommendation was to promote and bolster women’s networks of social support, and to include the involvement of partners in HIV risk reduction options. For heterosexual couples with potential unequal power dynamics, male involvement with women-centered prevention methods such as PrEP can improve adherence for optimal efficacy. Yet, in the absence of positive male reinforcement, community service providers must be the primary source of support and information for women who elect to choose such methods. To promote efficacy, an anonymous support group via phone could be used to provide information and support, while maintaining privacy. At a broader level, in the absence of family support, other key community leaders, including spiritual leaders, should be educated to lend credibility to and promote/support effective HIV prevention practices.

#### 5. Increase access to existing resources

There was a general belief regarding a need to increase awareness about community resources for HIV screening services and access to HIV care, such as Ryan White health coverage, which provides help for low-income people with HIV (Ryan White HIV/AIDS Program, 2022). The Haitian focus group felt that annual medical physicals, including HIV/STI screening, should be promoted—especially among youth. Individuals who receive annual medical check-ups or are actively engaged in care are more likely to have routine HIV/STI screenings (Cyrus et al., 2021). One benefit of identifying HIV infection prior to symptomatic presentation is that early detection can aid in the prevention of acute infection by sexual partners, and routine screening also presents opportunities to discuss HIV prevention options (Buchacz et al., 2005).

Latina women also acknowledged the need to promote existing programs that provided financial assistance for patients seeking PrEP, such as the Florida Department of Health PrEP campaign (Florida Department of Health, 2019), which subsidizes eligible individuals who lack insurance or are not able to afford PrEP or the Truvada® for PrEP medication assistance program, which assists eligible HIV-negative adults in the US (Gilead, 2018).

## IV. CONCLUSIONS

Increasing PrEP utilization among women of color requires a multi-tiered approach that addresses individual, structural, and community barriers while simultaneously drawing from the existing social networks and strengths of ethnic/community social groups and community organizations to facilitate the process. The Summit’s underlying theme, “we already have the tools to end the HIV epidemic,” brought awareness to current HIV prevention resources.

The challenge from the Summit was to address the factors hindering women’s empowerment to take control of their sexual health, starting with access to adequate care and PrEP use. As one participant commented, *when it comes to empowering women surrounding their sexual health, PrEP is viewed as the driving vehicle*. One of the Summit highlights was the importance of recognizing the multiplicity of sociocultural factors and their interaction and relationship to creating barriers for minoritized women accessing and using PrEP.

Improving communication skills with partners and providers will contribute to sustained women’s empowerment that can be translated into women protecting themselves sexually against HIV. Implicit in this approach is developing women’s ability to have the confidence to negotiate these relationships. One specific, practical exercise discussed in the focus groups was a recommendation that women prepare a list of questions before a doctor’s visit to ensure that all salient topics are covered during the medical encounter. As discussed in the recommendations, other approaches need to address social and systematic barriers to encourage environments that promote women taking an active role in their sexual health.

Reflections from the Summit emphasized cultural sensitivity and understanding in the provider-patient encounter (Bezreh et al., 2012; Laws et al., 2014). Cultural congruity between the provider and the client was highlighted as critical given that healthcare providers play a critical role in implementing PrEP in care settings. Recent studies have found that Latinos are the least likely to visit a doctor (Livingston et al., 2008; Machado, 2014). More than one-fourth of Latino adults in the United States lack a primary healthcare provider, almost 50% never visit a medical professional during the course of the year (Livingston et al., 2008), and they are more likely to delay care or drop out of treatment when symptoms disappear compared to their White counterparts (Machado, 2014). Data is limited for experiences with Haitian women, and further exploration is need to understand their perceptions and experiences with providers.

Healthcare providers that serve these populations may need to be encouraged and trained (when needed) to engage in non-judgmental, transparent conversations about sex and maintain patient confidentiality to foster trust. Providers can benefit from continuous education on recognizing personal biases, having greater consideration for patients’ needs, providing uniform care to all individuals, performing more detailed intakes without pressure or coercion and, finally, offering the integration of services to help meet basic needs and sexual healthcare. Another recommendation was to use allied health professionals, such as adherence counselor, to complement and support the services the primary care provider offers.

Cultural awareness was reinforced during the Summit and was deemed as essential by many group participants in developing and implementing successful interventions. Research has demonstrated that for prevention/intervention studies, cultural beliefs may affect communication and strategies for prevention, as well as attitudes regarding sexual health and contraception (Institute of Medicine Committee on Unintended, 1995). Various studies across the US among high-risk Latinos highlight several mediating factors between high-risk behaviors and acculturation: (a) low prevalence of sexual barrier use and HIV testing, (b) low acceptability of sexual barriers (male and female condoms), (c) the influence of poverty, violence, drug dealing, and an environment of crime, (d) perception of AIDS-related risk, and (e) and alcohol use (Levison et al., 2018). Although the perceived risk of HIV infection among Latina women may be lower than it should be because of misinformation, avoidance of HIV testing may occur because of fear of stigmatization with traditional support systems for bringing “shame” on their family and their community by determining their HIV status (Villar-Loubet et al., 2011). In this sense, the power of stigma cannot be understated; stigma discourages women at risk for HIV from testing or seeking HIV prevention tools and prevents them from speaking openly with their sex partners about safer sex options.

There are still many gaps with respect to PrEP and transgender populations. In retrospect, although important points were raised in the three discussion groups concerning transgender women’s experiences, the Summit would have benefited from a separate group for transgender women. Moving forward on the issues regarding transgender-specific needs, additional formative work is being completed in a follow-up study to explore more transgender-specific barriers and facilitators of PrEP use, including substance use and other social determinants (Kiplagat et al., 2020).

In summary, the recommendations and suggestions presented in this article represent the thoughtful assessments of community members, health practitioners, scientists, and government officials who have dedicated years to HIV prevention and women’s health. This article synthesizes and reflects the wealth of information discussed at this progressive forward-looking Summit. It is the desire of the authors that this article encourages further discussion and contributes to the conversation that will improve practices and interventions to advance the wellbeing and empowerment of all minoritized women globally. The objective moving forward is to build on these recommendations to develop action-oriented agendas that integrate research, practice, and policy, with the goal of empowering African American, Latina, and Haitian *women to take control of their sexual health*.

## Data Availability

All data produced in the present study are available upon reasonable request to the authors

